# Effects of Resistance Training Combined with a Ketogenic Diet: A Systematic Review and Meta-Analysis

**DOI:** 10.1101/2024.06.19.24308878

**Authors:** Luis Roberto Sinott, Clédia Silveira Flores da Silva, Alice Künzgen Scheer, Amanda Barbosa Atrib, Augusto Scheneider, Carlos Castilho Barros

**Affiliations:** Nutrigenomic Laboratory, Universidade Federal de Pelotas, Pelotas, Brasil

**Keywords:** high-fat diet, carbohydrate-restricted diet, body composition, physical activity, muscle hypertrophy

## Abstract

Weight loss treatments require adherence to physical exercise and diet. Restrictive diets have been proposed for obesity treatment, including a ketogenic diet that are high in lipids, moderate in proteins, and low in carbohydrates. In recent years, there has been criticism of this diet because of the reduction in fat-free mass and, consequently, a reduction in basal energy expenditure, which is considered negative in obesity treatment. However, resistance training is known to promote skeletal muscle hypertrophy. The hypothesis for this review was: “Resistance training is sufficient to maintain lean mass during diets that cause ketosis.” Despite the slight reduction in lean mass identified in the meta-analysis, some authors reported no loss in physical performance. Others suggested that this difference in lean mass is associated with water loss in the participants, which aligns with a few studies that reported a final phase with carbohydrate reintroduction into the diet. Our results indicated physical exercise was an important tool for maintaining lean mass in individuals who consumed carbohydrate-restricted diets that cause ketosis.

## Introduction

Resistance training is primarily used to gain muscle mass through anaerobic stimulation of muscle hypertrophy (SCHOENFELD et al. 2019). Resistance exercise can also reduce blood pressure (DE SOUSA et al. 2017) and blood glucose levels (TAKENAMI et al. 2019), increase basal metabolic rate (BMR) (DOLEZAL; POTTEIGER 1998), assist in weight loss (GOLDFIELD et al. 2017) and reduce cardiovascular mortality risk (SAEIDIFARD et al. 2019). The increase in total energy expenditure achieved through resistance training and BMR increase are consequences of the metabolic effects from exercise, lean mass gain (ARISTIZABAL et al. 2015; MACKENZIE-SHALDERS et al. 2020), and weight loss (HUNTER et al. 2015; MACKENZIE-SHALDERS et al. 2020).

Weight loss treatments primarily involve a combination of diet and physical exercise (MOZAFFARIAN et al. 2011). Although some professionals contest these restrictive strategies, weight loss diets tend to be constraining (VAN HORN 2014). Dietary restrictions can be based on energy deficits (DE SOUZA et al. 2012) or macronutrient composition (BUENO et al. 2013). Thus, they can include conventional hypocaloric (low energy density) (FOCK; KHOO 2013), low-fat (RAZAVI ZADE et al. 2016), low-carbohydrate (CARE; SUPPL 2020; KELLY; UNWIN; FINUCANE 2020), or high-protein diets (ZHAO et al. 2018).

Among the restrictive diets, the ketogenic diet restricts carbohydrate intake without necessarily restricting the total energy intake (KANG et al. 2020). A ketogenic diet is high in lipids, moderate in proteins, and low in carbohydrates (GIUGLIANO et al. 2018). Carbohydrate intake is typically limited to 50 g/day (MUSCOGIURI et al. 2019) or less than 10% of the total energy value, stimulating the production of ketone bodies that can be used as an energy source to maintain metabolism, although they are also excreted in the urine. Physiological ketosis promotes rapid initial weight loss due to increased diuresis in response to reduced glycogen levels in the tissues. Metabolic changes can occur due to the lack of carbohydrates in the diet or hormonal changes, notably the reduction of insulin release by the pancreas and the consequent reduction of gluconeogenesis stimulation, which requires energy expenditure. After stabilizing water loss, weight reduction can remain constant if the total energy intake is kept below the energy expenditure and other factors, such as metabolic waste and the elimination of ketone bodies in the urine, are considered. However, this depends on an individual’s adaptive process, as few people can endure the adaptation phase or maintain this diet for long periods (MUSCOGIURI et al. 2019). Although a ketogenic diet has several benefits, it can cause adverse effects such as dehydration, hypoglycemia, lethargy, halitosis, and gastrointestinal disturbances (O’NEILL; RAGGI 2020). These side effects are generally moderate and can be overcome with monitoring and adjustments in fiber and vegetable intake.

In recent years, diets that cause ketosis have been criticized because of their tendency to reduce lean mass (fat-free mass [FFM]) and, as a consequence, reduce basal energy expenditure, which is considered a negative effect in obesity treatment.

This study hypothesized that resistance training is an important tool for maintaining FFM during a ketogenic diet. Thus, we reviewed the literature and compiled articles that combined a ketogenic diet with resistance training; thereby conducted a systematic review and meta-analysis using their contents.

## Methodology

The preestablished protocol was registered on the PROSPERO website (CRD42018116655). The Preferred Reporting Items for Systematic Reviews and Meta-Analyses (PRISMA) guidelines were followed (LIBERATI et al. 2009). To meet the eligibility criteria, articles were required to include experimental studies on ketogenic diets and resistance exercise in humans or animals. A systematic search of scientific works was conducted using three databases (PUBMED, Embase, and SportDiscus) published until June 10, 2020. The search algorithm is described in Supplementary Material (Table S1). In general, the search used the term “OR” between words identifying ketogenic diets and words identifying resistance exercises, and “AND” between the two groups. Adjustments were made to the algorithms for different databases. Duplicate articles, non-English or Portuguese articles, reviews, incomplete articles with only abstracts, and case studies were excluded. The remaining articles were filtered using the following inclusion and exclusion criteria:

### Inclusion criteria

randomized controlled clinical trials that evaluated resistance training sessions associated with a ketogenic diet in men or women with or without chronic diseases, trained or untrained, aged >18 years. The diets had to include <50 g of carbohydrates/day (up to 10% total caloric value or confirmed ketosis by circulating β-hydroxybutyrate (βHB) ≥0.5 mmol/L. The resistance training and diet had to be applied for at least 4 weeks, with body composition data read at the start and end of the study.

### Exclusion criteria

Articles that did not report ketosis results for the participants, did not present lean mass values, had treatment durations of less than 4 weeks, or lacked a control group were excluded.

Two authors selected the articles. Group sizes, FFM changes, and standard deviations were extracted for the meta-analysis. When data were presented exclusively in graphs, GetData Graph Digitizer was used to extract them. The meta-analysis was performed using STATA software and the random method. The selected studies underwent bias evaluation using the RoB 2.0 tool (The Cochrane Collaboration, 2019).

A total of 567 articles 295 in EMBASE, 199 in PubMed, and 73 in SPORTDiscus. Of these, 92 duplicates were excluded. Studies in which body composition data were collected only at the beginning or end of the experiment were also excluded (Figure 1). When data were missing from the manuscript, the corresponding authors were contacted.

**Figure 1:**
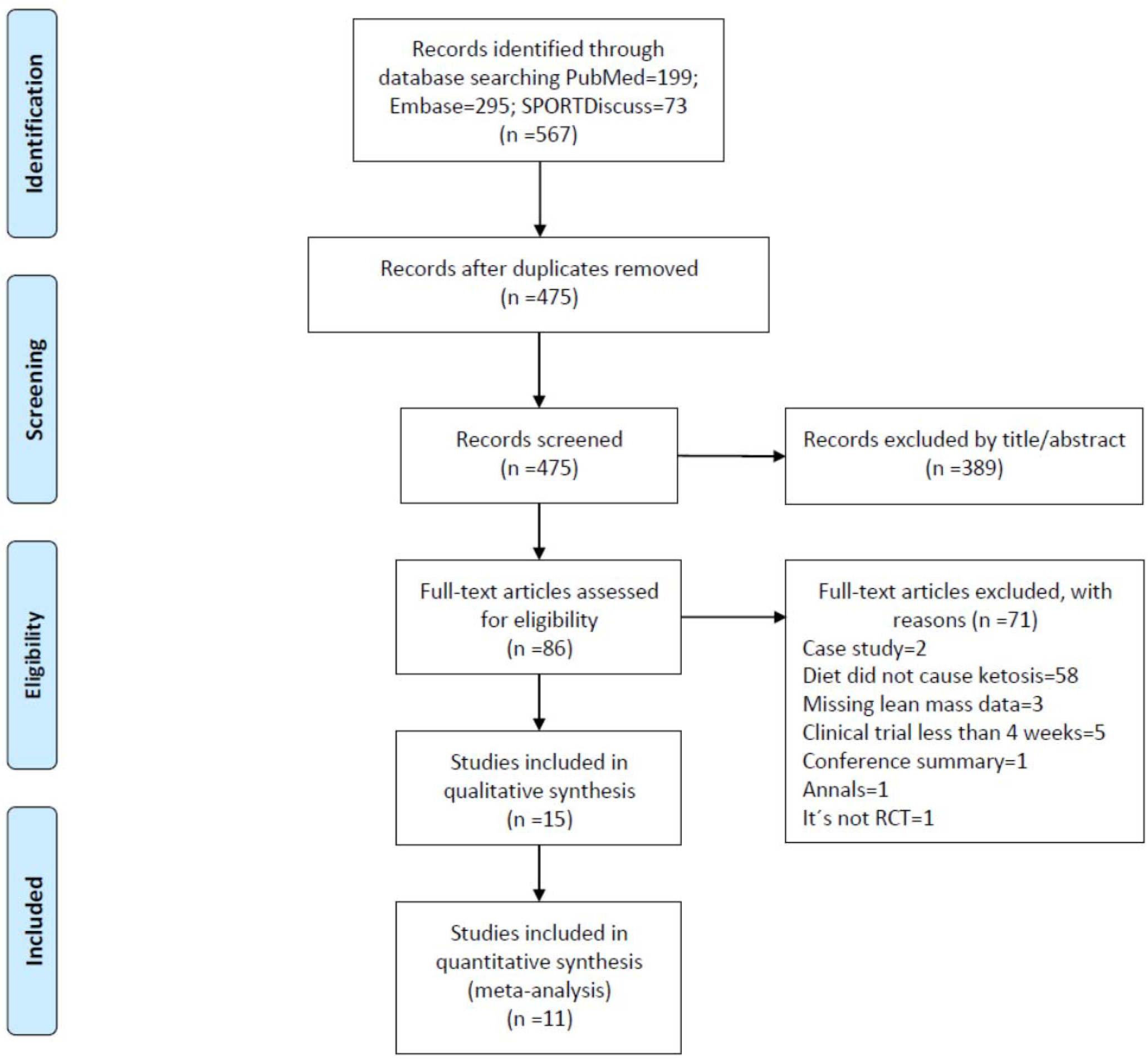
Flowchart of the article selection process.

## Results and Discussion

### Effect of Resistance Exercise on Lean Mass Preservation in Humans

This study initially intended to verify whether resistance exercise was an effective tool for maintaining FFM during weight loss treatment with a ketogenic diet. Studies with experimental designs allowing the comparison of participants who consumed a ketogenic diet with and without resistance exercise were sought. However, such studies were not found in the literature. Therefore, studies presenting only exercise training groups consuming different diets were used to compare the FFM changes among diets.

Ten studies were selected for the meta-analysis. The details of these studies, including the sample population, dietary interventions, exercise protocols, sample size, intervention duration, and main results, are presented in Table 1. Despite the heterogeneity of the studies, the meta-analysis indicated a slight reduction of FFM in participants who consumed a ketogenic diet (Figure 2). The effect represented by the pooled D+L of SMD was -0.347 with a 95% confidence interval between -0.549 and -0.144. This analysis showed 2948 (df =10) p=0.001, I-squared (variation in SMD attributable to heterogeneity) = 66.1%, and between-study variance estimate tau-squared = 0.1252; SMD=0: z=3.36 p<0.001. The detailed results of the analysis are presented in Table 2.

**Table 1.** Selected Studies, Dietary Interventions, Exercise Protocols, and Main Results.

| Study | Population (n) | Diets | Exercise Protocol | Main Results |
| --- | --- | --- | --- | --- |
| Perissiou et al. (2020) | 64 male and female participants with obesity | Groups: low-carbohydrate meals and control (standard dietary counseling) | Both groups had a structured supervised exercise program for 8 weeks | Short-term low-carb diet combined with prescribed exercise was associated with greater muscle mass loss compared to the group that received standard dietary counseling and supervised exercise. |
| Vargas-Molina (2020) | 21 women practicing strength exercise | Standard diet group (n=11) and ketogenic diet group (n=10) | 4 training sessions per week (divided into 2 cycles of 4 weeks) during 8 weeks. | The ketogenic diet helped reduce fat mass and maintain FFM after 8 weeks of resistance training in trained women. |
| Wilson et al. (2020) | 25 college men | Groups: ketogenic diet and standard diet for 10 weeks with carbohydrate reintroduction in weeks 10 to 11 | Resistance training program | Lean mass increased in both groups in week 10. However, only the ketogenic diet group increased lean mass between weeks 10 and 11 (carbohydrate reintroduction) and fat mass decreased in both groups. The ketogenic diet was used in combination with resistance training to cause favorable changes in body composition. |
| Greene et al. (2018) | Intermediate competitive lifters | Ad libitum diets: usual diet (250 g carbohydrates) and ketogenic diet (50 g or 10% VCT) = 3-month crossover | Habitual training during both diet phases | Lean mass changes: -226 SE 064; 95% CI -410 to -042. No reduction in physical performance. |
| McSwiney et al. (2018) | 20 trained male athletes | Groups: high carbohydrate and low carb | Both groups performed the same training intervention (resistance strength and high-intensity interval training ) = 12 weeks of diet and training | The low-carb group during the 12-week keto-adaptation and physical training period improved body composition, fat oxidation during exercise, and specific performance measures relevant to competitive endurance athletes. |
| Vargas (2018) | 24 healthy men | Ketogenic diet group (n=9); non-ketogenic diet group (n=10) and control group (n=5) | 4 sessions per week of a hypertrophy training protocol: 2 upper limb days and 2 lower limb days with 72 hours rest between sessions; 8-week duration | No significant increases in total body weight and muscle mass were observed in the ketogenic diet group. The non-ketogenic diet group had increases in these parameters. The control group showed no changes in body weight or lean mass. |
| Wood(2012) | 32 men | Groups: low fat; low fat and progressive resistance exercise ; carbohydrate-restricted diets and carbohydrate-restricted diets with progressive resistance exercise | Supervised strength training 3 times a week = 12 weeks | The percentages of appendicular FFM loss were 2.75%, 1.59%, 1.57%, and 1.73%, respectively, for the 3 groups. Possibility to compare carbohydrate-restricted diet with and without exercise. |
| Jabekk(2010) | 18 untrained women with a BMI ≥ 25 kg/m<sup>2</sup> | Groups: standard diet, low carb diet, and ketogenic diet | 60–100 minutes of varied resistance exercise twice a week = 10 weeks | Resistance exercise combined with a ketogenic diet reduced body fat without significantly altering lean mass. |

**Table 2.** Meta-Analysis Results.

| Study | SMD | [95% Conf. Interval] |  | % Weight |
| --- | --- | --- | --- | --- |
| Greene, D.A. 2018 | -2.858 | -4.022 | -1.695 | 3.03 |
| Kerksick, C.M.; 2010 | 0.000 | -0.397 | 0.397 | 25.98 |
| Kerksick, C.M.; 2010 | -0.126 | -1.057 | 0.806 | 4.72 |
| Tay, J.; 2015 | -0.060 | -0.505 | 0.384 | 20.72 |
| Vargas-Molina, S.; 2020 | 0.000 | -0.856 | 0.856 | 5.58 |
| Vargas, S.; 2018 | -0.906 | -1.856 | 0.045 | 4.53 |
| McSwiney, F.T.; 2018 | 0.027 | -0.854 | 0.908 | 5.28 |
| Wood, R.J.; 2012 | -0.164 | -1.154 | 0.825 | 4.18 |
| Jabekk, P.T.; 2010 | -0.857 | -1.887 | 0.173 | 3.86 |
| Perissiou, M.; 2020 | -0.735 | -1.242 | -0.228 | 15.93 |
| Wilson, J.M.; 2020 | -0.731 | -1.544 | 0.081 | 6.20 |
| I-V pooled SMD | -0.347 | -0.549 | -0.144 | 100.00 |

**Figure 2:**
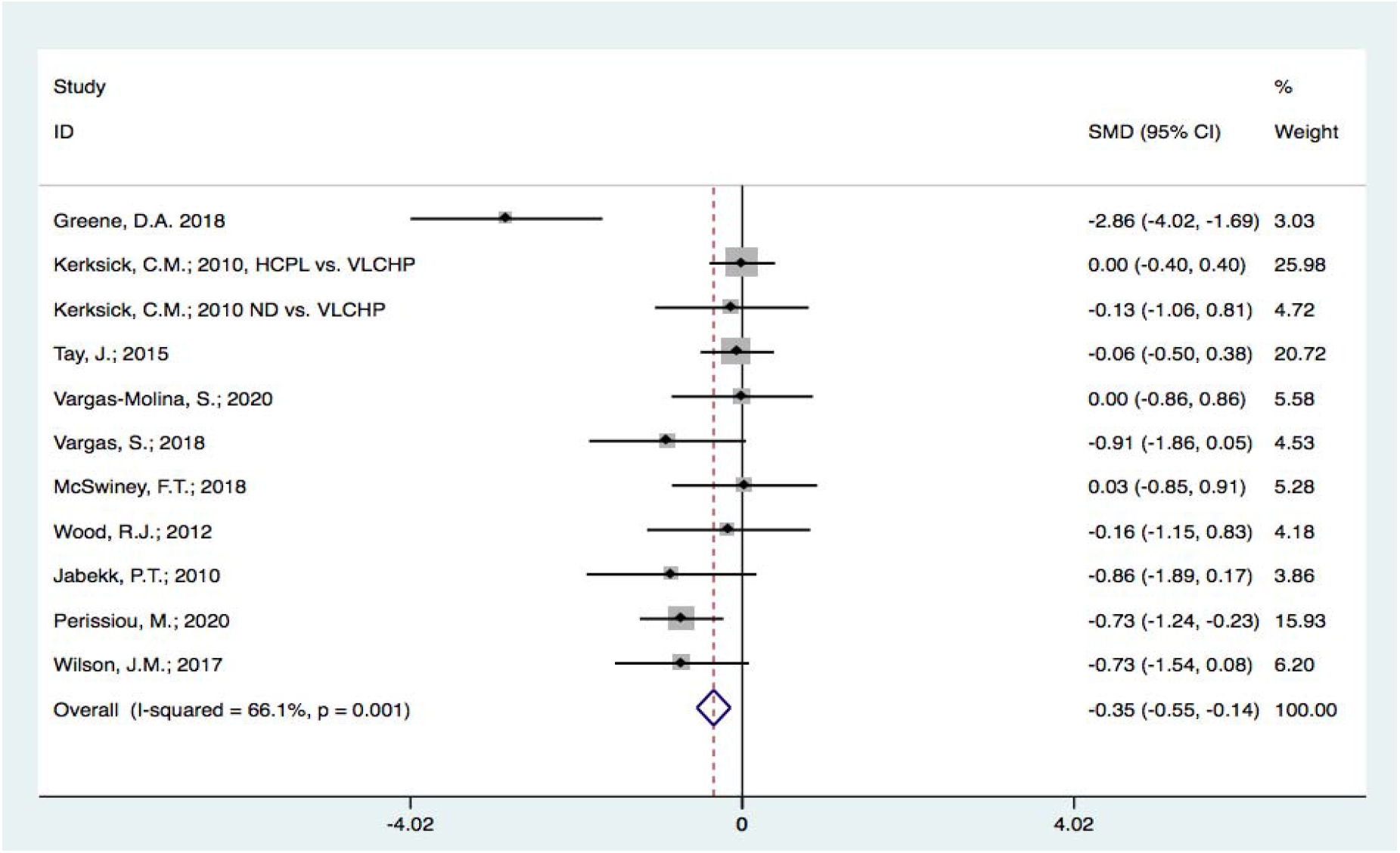
Forest plot graph showing heterogeneity among the studies in the meta-analysis and a small reduction in FFM among the patients in the analyzed studies. Chi-square heterogeneity = 29.48 (df = 10), p = 0.001; I-squared (variation in SMD attributable to heterogeneity) = 66.1%; between-study variance estimate T-squared = 0.1252; SMD = 0: z = 3.36, p < 0.001.

### Effect of Diets on Body Composition and Physical Performance

Given the results obtained in the meta-analysis, we examined how the authors discussed the causes of the reduction in FFM observed in the studies and whether this loss was reflected in decreased sports performance. Table 3 summarizes the effects observed in the 10 selected articles, highlighting whether carbohydrates were reintroduced at the end of the study, a FFM reduction was observed, the authors attributed this decrease to water loss due to the low-carbohydrate nature of the diets, athletic performance was reduced, and a ketogenic diet had a greater effect on reducing body weight and fat mass than the control diet.

**Table 3:** Selected Studies and Their Effects Regarding Carbohydrate Reintroduction and Changes in Body Composition.

| **Study** | **Carbohydrate Reintroduction** | **Reduction in FFM** | **Blame Water\*** | **Performance** | **Body Weight** | **Fat Mass** |
| --- | --- | --- | --- | --- | --- | --- |
| Greene 2018 | No | Yes | Yes | Not affected | Reduced | Reduced |
| Kerksick 2010 | Yes | Very small | No | Not evaluated | Reduced | Reduced |
| Tay 2015 | No | No | N/A | Not evaluated | Similar | Similar |
| Vargas-Molina2020 | No | No | N/A | Not affected | Reduced | Reduced |
| Vargas 2018 | No | No difference inside group, but reduced comparing NKD group | Do not discuss water content | Not evaluated | Reduced | Reduced |
| McSwiney 2018 | No | No | N/A | Not affected | Reduced | Reduced |
| Wood 2012 | No | No | N/A | Not affected | Reduced | Reduced |
| Jabekk 2010 | No | No difference inside group, but reduced comparing groups | Do not discuss water content | Not evaluated | Reduced | Reduced |
| Perissiou 2020 | No | Yes | Yes | Not evaluated | Reduced | Similar reduction |
| Wilson 2020 | Yes | Increase muscle in KD | N/A | Increased performance | Similar after reload carbo | Similar |

\*Description if the authors commented on the loss of water to justify the differences in FFM.
N/A, not applicable because there was no FFM reduction.

Two studies that reintroduced carbohydrates into the diet at the end of the experiment for at least 1 week reported a small loss or a gain in FFM after carbohydrate reintroduction (KERKSICK et al., 2010; WILSON et al., 2020). Two other studies reported no change in FFM before and after a ketogenic diet despite a difference from the control diet, which showed an increase in FFM after resistance training (VARGAS-MOLINA et al., 2020). Four other studies reported no significant differences in FFM between the control and ketogenic diet groups (VARGAS-MOLINA et al., 2020; MCSWINEY et al., 2018; TAY et al., 2015; WOOD et al., 2012). Of the studies that observed a significant reduction in FFM, both discussed in their results that this effect could have been associated with water loss, a well-known phenomenon caused by diuresis in patients on carbohydrate-restricted diets (PERISSIOU M, BORKOLES E, KOBAYASHI K, 2020; GREENE DA, VARLEY BJ, HARTWIG TB, CHAPMAN P, 2018).

Most studies reported that the ketogenic diet groups experienced a reduction in FFM; however in one of the two studies with carbohydrate reintroduction at the end this difference was reversed (WILSON et al., 2020).

### Proposed Mechanisms for Lean Mass Maintenance through Exercise

Among the selected studies, there was evidence related to the mechanisms involved in the effects of the ketogenic diet and resistance exercise on FFM variations. Since the activation of the AKT pathway has often been associated with muscle hypertrophy, a reduction in insulin release in low-carbohydrate diets might be the main cause of muscle mass loss. Although this hypothesis seems evident, studies in humans and animal models have failed to demonstrate this process, and more data are needed to verify this connection. Other compensatory mechanisms are also stimulated by exercise. A study in rats showed that basal levels of (p-rps6, p-4EBP1, and p-AMPKα) were similar between diets, although serum insulin, serum glucose, and various essential amino acids levels were lower in rats fed a low-carb ketogenic diet (LCKD). Additionally, LCKD-and standard diet-fed rats exhibited increased levels of post-exercise muscle protein synthesis; however, no dietary effects were observed. Animals fed these diets and trained exhibited similar increases in relative hindlimb muscle mass compared to their sedentary counterparts, although there was no difference between the diet groups (ROBERTS et al., 2016).

A review (PAOLI et al., 2019) suggested that the overloaded regulatory systems controlling muscle hypertrophy as an effect of exercise reflect alterations in protein synthesis and degradation and consequent muscle hypertrophy. They also suggested that carbohydrate restriction in the diet involves the reduction of some hormones and their signaling, concluding that the data provided in the scientific literature suggest only a minor or no effect from the ketogenic diet on muscle mass with concurrent resistance training. The results of the present study support this hypothesis.

### Conclusion

This study systematically reviewed the literature to verify whether resistance training was an effective tool for maintaining FFM in individuals undergoing carbohydrate-restricted diets to induce ketosis. Although few studies have compared groups of ketogenic diets with or without exercise, we were able to compare individuals who underwent resistance training with or without ketosis-inducing diets. The meta-analysis indicated a slight reduction of FFM in individuals who consumed ketogenic diets, a reduction that was reversed by carbohydrate reintroduction, or described as a possible effect of water loss due to the low-carbohydrate nature of the diets. In addition, to date, no study has demonstrated a loss of muscle strength associated with FFM loss or a ketogenic diet. These results strongly suggest that resistance training is an efficient tool for preserving lean mass in patients who consume diets that induce ketosis. Clinical trials comparing individuals with or without exercise and those consuming carbohydrate-restricted diets are necessary to confirm this conclusion.

## Data Availability

All data produced in the present work are contained in the manuscript

