## Supplementary material for "Effects of Resistance Training Combined with a Ketogenic Diet: A Systematic Review and Meta-Analysis": Table S1

Table S1: Search algorithm and data bases.

| DATA BASE | SEARCH ALGORITHM |
| --- | --- |
| PUBMED | (((((((((((((((((((((“Diet, Ketogenic”[MeSH]) OR “Diet, Ketogenic”) OR “Ketogenic Diet”) OR “Diets, Ketogenic”) OR “Ketogenic Diets”) OR “Diet, Carbohydrate-Restricted”[MeSH]) OR “Diet, Carbohydrate-Restricted”) OR “Diet, Carbohydrate Restricted”) OR “Diet, Low Carbohydrate”) OR “Carbohydrate Diet, Low”) OR “Carbohydrate Diets, Low”) OR “Diets, Low Carbohydrate”) OR “Low Carbohydrate Diets”) OR “Carbohydrate-Restricted Diet”) OR “Carbohydrate Restricted Diet”) OR “Carbohydrate-Restricted Diets”) OR “Diets, Carbohydrate-Restricted”) OR “Low-Carbohydrate Diet”) OR “Diet, Low-Carbohydrate”) OR “Diets, Low-Carbohydrate”) OR “Low Carbohydrate Diet”) OR “Low-Carbohydrate Diets”)) AND ((((((((((((((((((((((“Resistance Training”[MeSH]) OR “Resistance Training”) OR “Training, Resistance”) OR ""Strength Training”) OR “Training, Strength”) OR “Weight-Lifting Strengthening Program”) OR “Strengthening Program, Weight-Lifting”) OR “Strengthening Programs, Weight-Lifting"") OR “Weight Lifting Strengthening Program”) OR “Weight-Lifting Strengthening Programs"") OR “Weight-Lifting Exercise Program”) OR “Exercise Program, Weight-Lifting"") OR “Exercise Programs, Weight-Lifting”) OR “Weight Lifting Exercise Program”) OR “Weight-Lifting Exercise Programs"") OR “Weight-Bearing Strengthening Program”) OR “Strengthening Program, Weight-Bearing”) OR “Strengthening Programs, Weight-Bearing”) OR “Weight Bearing Strengthening Program”) OR “Weight-Bearing Strengthening Programs”) OR “Weight-Bearing Exercise Program”) OR “Exercise Program, Weight-Bearing”) OR “Exercise Programs, Weight-Bearing”) OR “Weight Bearing Exercise Program”) OR “Weight-Bearing Exercise Programs”)) |
| SPORTDiscus | ( SU ( ("RESISTANCE training" OR "WEIGHT training" OR "STRENGTH training" OR "WEIGHT lifting") ) OR TX ( (“resistance exercise” OR “resistance exercise training” OR “weight bearing exercise” OR “load carrying” OR “weight bearing” OR “weight-bearing”) ) ) AND ( SU ( (“KETOGENIC diet” OR “LOW-carbohydrate diet” OR “HIGH-protein diet”) ) OR TX ( (“High Protein Carbohydrate Restricted Diet*” OR “High-Protein Carbohydrate-Restricted Diet*" OR “Low Carbohydrate High Protein Diet*” OR “Atkins Diet” OR “South Beach Diet” OR “keto diet” OR “ketotic diet” OR “ketogenous diet” OR “hp-lc diet” OR “hplc diet” OR “atkin diet”) ) ) |
